# Circular functional analysis of OCT data for precise identification of structural phenotypes in the eye

**DOI:** 10.1101/2021.02.07.21251275

**Authors:** Md. Hasnat Ali, Brian Wainwright, Alexander Petersen, Ganesh B. Jonnadula, Meghana Aruru, Harsha L. Rao, M. B. Srinivas, S. Rao Jammalamadaka, Sirisha Senthil, Saumyadipta Pyne

## Abstract

Progressive optic neuropathies such as glaucoma are major causes of blindness globally. Multiple sources of subjectivity and analytical challenges are often encountered by the clinicians in the process of early diagnosis and clinical management of these diseases. In glaucoma, the structural damage is often characterized by neuroretinal rim (NRR) thinning of the optic nerve head, and other clinical parameters. Optical coherence tomography (OCT) is a popular and quantitative eye imaging platform for precise and reproducible measurement of such parameters in the clinic.

Baseline structural heterogeneity in the eyes can play a key role in the progression of optic neuropathies, and thus present challenges to clinical decision-making. To address this, large and diverse normative OCT databases with mathematically precise description of phenotypes can help with early detection and characterization of the different phenotypes that are encountered in the clinic. In this study, we generated a new large dataset of OCT generated high-resolution circular data on NRR phenotypes, along with other clinical covariates, of nearly 4,000 healthy eyes as part of a well-established clinical cohort (LVPEI-GLEAMS) of Asian Indian participants.

In this study, we (1) generated high-resolution circular OCT measurements of NRR thickness in a given eye, (2) introduced CIFU, a new computational pipeline for <u>CI</u>rcular <u>FU</u>nctional data modeling and analysis that is demonstrated using the OCT dataset, and (3) addressed the disparity of representation of the Asian Indian population in normative OCT databases. We demonstrated CIFU by unsupervised circular functional clustering of the OCT NRR data, meta-clustering to characterize the clustering output using clinical covariates, and presenting a circular visualization of the results. Upon stratification by age, we identified a healthy NRR phenotype cluster in the age group 40-49 years with predictive potential for glaucoma.

## 1. Introduction

Progressive optic neuropathies such as glaucoma can cause irreversible blindness, especially when left untreated or diagnosed late. Indeed, early detection and management hold the key to slowing the progressive loss of vision and preventing blindness due to many chronic and age-related degenerative eye diseases. Glaucoma, for instance, is the second-leading cause of blindness worldwide^1^. In 2020, an estimated 80 million individuals worldwide had glaucoma and this number is expected to increase to over 111 million by 2040^2^.

There are multiple sources of subjectivity and analytical challenges that are often encountered by the clinicians in the process of early diagnosis and clinical management of these diseases. In glaucoma, the functional damage is established most commonly by the occurrence of visual field (VF) loss whereas the structural damage is often characterized by neuroretinal rim (NRR) thinning of the optic nerve head (ONH), and loss of retinal nerve fibers, which are the axons of retinal ganglion cells (RGC). On the functional side, while standard automated perimetry (SAP) has been the gold standard for detection of VF loss, often 30% of RGC loss may have already occurred before VF defects could be detected by SAP^3^.

On the structural side, biological heterogeneity of ONH phenotypes, with or without any neuropathy, can present challenges to clinical decision-making. For instance, NRR area has been found to normally decline at the rate of 0.28%-0.39% per year^4^. There is no single, specific management guidance for patients with diverse morphology of ONH^5^. For instance, to assess the progression of glaucoma, one of the parameters assessed is the optic cup-to-disc ratio (CDR) which is calculated by comparing the diameter of the “cup” portion of the optic disc with the total diameter of the latter. Yet, while a large CDR may indicate glaucoma or other pathology, deep yet stable (over age) cupping, i.e., a normal physiologically large optic disc cup, can occur due to genetic factors in the absence of any disease or associated clinical covariates (e.g., high intraocular pressure)^6^. It is of great importance that sources of natural variation are rigorously understood thereby controlling for subjectivity in diagnosis.

In the clinic, non-invasive, high-resolution eye imaging platforms such as spectral-domain optical coherence tomography (OCT) provide excellent glaucoma diagnostic performance, especially in early stages of disease^7,8^. The quantitative and reproducible OCT data provide objective measurement of ONH parameters, NRR area, retinal nerve fiber layer (RNFL), macular thickness, etc., which are used by clinicians to identify structural damage. For instance, Zeiss Cirrus HD-OCT platform uses the clinically invisible but OCT detectable Bruch’s membrane opening (BMO) as the landmark to measure the amount of NRR tissue in the optic nerve. It has been reported that NRR thickness calculation by Cirrus HD-OCT has high reproducibility and glaucoma diagnostic ability, and a low rate of incorrect optic disc margin detection^9–11^. The platform generates a comparative report of a patient’s data based on its normative database.

The OCT platform performs circular scans of the eye which, as in many biomedical technologies, are examples of measurements that are recorded or indexed at different directions, say, at given angular positions around a central point. Unlike the analyses of “linear” data points that reside on the real line or Euclidean spaces, directional data requires special and altogether different treatment. For instance, a direction in two-dimensional plane can be represented as a point on the circumference of a unit circle, or simply as an angle, but neither representation is unique, as both depend on the selection of some appropriate “zero-direction” from which to start measuring, as well as the sense of rotation, viz., clockwise or anti-clockwise. The unique properties of circular data – for instance, if one wishes to compare two such scans with a distance measure – are appropriately addressed by the field of circular statistics^12^.

Traditional OCT analysis may involve division of the circle around ONH into 4 fixed quadrants, or 12 clock-hours, to record measurements at these sectors. In this study, we extended such data collection to divide the same circle (of total 360 degrees) into much finer segments of 2 degrees each, and thus, generate 180 circular data points for each clinical sample (human eye). These rich and evenly spaced high-resolution circular (HRC) data allows for natural application of functional data analysis (FDA) where the data are not viewed as points but as curves or mathematical functions. Not to be confused with alternate usage of the term “function” (such as in vision), FDA is increasingly popular in biomedical informatics due to the emergence of new monitoring technologies that can record data as curves^13–15^. The HRC OCT data for each sample can be modeled as a circular function or curve, with angle and magnitude being the independent and dependent variables, respectively. In this study, we describe methods for novel circular functional analysis of OCT data, and apply it for unsupervised identification of NRR phenotypes in healthy eyes.

Progressive and degenerative eye diseases benefit from pre-existing knowledge and cumulative collection and description of normal phenotypes as these may help to identify early and characterize precisely the new phenotypes that emerge over time. While some normative OCT databases do exist, they are generally limited in their size and diversity. Thus, the breakup of their ethnic representation may not reflect the actual epidemiologic distribution of the disease. For instance, only 1% of the popular Cirrus HD-OCT platform’s normative database is of Asian Indian origin^16^, although India contributes to more than 12% of the global cases of both primary open-angle and primary angle-closure glaucomas^17^. Towards this, we generated a new large HRC OCT dataset on NRR phenotypes, along with other clinical covariates, of 3973 healthy eyes as part of a well-established clinical cohort (LVPEI-GLEAMS) at the L.V. Prasad Eye Institute, Hyderabad, India.

The main objectives of the present study are to: (1) generate HRC OCT data in the form of 180 circular measurements of NRR thickness in a given eye, (2) introduce CIFU, a computational pipeline for <u>CI</u>rcular <u>FU</u>nctional data modeling and analysis that is demonstrated using the OCT dataset, and (3) address the disparity of representation of the Asian Indian population in normative OCT databases. In the next section, we describe the clinical cohort and the protocol for data generation as well as the algorithm of CIFU for unsupervised circular functional clustering of the OCT NRR data, followed by meta-clustering to characterize the clustering output using clinical covariates of glaucoma. In the following section, the results of CIFU analysis are described with help of circular visualization. In particular, upon stratification of the samples by age, we identified a healthy NRR phenotype cluster in the age group 40-49 years, and having the highest mean values of cup volume and average CDR among all clusters, with predictive potential for glaucoma. We end with discussion of the CIFU approach and its potential applications to future work.

## 2. Data and Methods

### 2.1. Data

All participants were selected from a population-based study LVPEI-GLEAMS (LVPEI Glaucoma Epidemiology And Molecular Genetic Study^17^) conducted by the L.V. Prasad Eye Institute (LVPEI), Hyderabad, India. Data were collected on a total of 3973 (OD:1981; OS:1992) healthy eyes of 2222 participants from the southern Indian state of Andhra Pradesh, India. Written informed consent was obtained from all participants to participate in the study, and the ethics and review committee of the LVPEI reviewed and approved the methodology and was conducted in strict adherence to the tenets of the Declaration of Helsinki. The inclusion criteria used were age >= 40 years, best-corrected visual acuity of 20/40 or better, spherical equivalent of ±6 Diopters, good quality stereo optic disc photographs, and no media opacities (signal strength >=6). The exclusion criteria used were intraocular surgery within the previous 6 months, and any retinal (including macular) or neurologic diseases other that could confound the structural measurements with SD-OCT.

Healthy eyes were defined by the absence of anterior and posterior pathology. Each digital optic disc photograph was evaluated by three glaucoma specialists independently. The specialists were masked to the other clinical findings and the other imaging outcomes of the subjects. Eyes were excluded from the study in case of any disagreements among the specialists. All participants underwent a comprehensive ophthalmic examination which included detailed medical and systemic history. The means of clinical determination included best-corrected visual acuity measurement, slit-lamp photographs (Topcon, Bauer Drive, Oakland, NJ), Goldmann applanation tonometry (Hagg-Streit AT 900, Hagg-Streit AG, Switzerland), gonioscopy with a Sussman four mirror gonioscope (Volk Optical Inc, Mentor, Ohio, USA), dilated fundus examination, central corneal thickness (CCT) assessment, Humphrey visual fields (HVF) with 24-2 Swedish Interactive threshold algorithm (SITA; Carl Zeiss Meditec Inc. Dublin, CA). Visual fields (VF) were considered if false positive, false negative, and fixation losses were less than 20%, and all the stereophotographs of the optic disc had good quality.

In addition, digital optic disc photography and spectral-domain optical coherence tomography imaging with Cirrus HD-OCT (software version 9.0.0.281; Carl Zeiss Meditec, Dublin, CA, USA) were used. This is a computerized instrument that acquires and analyzes cross-sectional and three-dimensional tomograms of the eye using spectral-domain optical coherence tomography (SD-OCT) technology. The algorithm of the instrument automatically identifies the optic disc margin as the termination of Bruch’s membrane (BM). BM opening (BMO) is used as the landmark to measure the amount of neuro-retinal rim tissue in the optic nerve. Optic Disc Cube 200×200 protocol was used to scan the ONH and peripapillary area through a 6 mm square grid, which consists of 200 horizontal linear B-scans and each composed of 200 A-scans. First, the Cirrus HD-OCT algorithm identifies the center of ONH and then automatically places a calculation circle of 3.46 mm diameter evenly around it. The circular scan starts at an extreme temporal point and moves around the ring in the superior direction, then nasal, then inferior, then back to temporal (TSNIT). The circular measurements are made clockwise for the right eye and counter-clockwise for the left eye. NRR thickness is measured by the amount of neuro-retinal tissue in the optic nerve around the entire edge of the optic disc. Zeiss Cirrus HD-OCT used Bruch’s membrane opening– minimum rim width (BMO-MRW) to measure the rim area. The BMO-MRW is the shortest distance from BMO to the retinal internal limiting membrane. The advanced export functionality was used to record the NRR thickness values at 180 points in TSNIT order spaced evenly by 2 degrees (from 2°–360°) around the circle. We refer to this as our NRR OCT high-resolution circular (HRC) data. The data were stratified into 3 age groups: (1) 40-49 years, (2) 50-59 years, and (3) 60 years and older.

### 2.2. Methods

We describe CIFU pipeline for circular functional modeling and clustering of HRC OCT data, followed by metaclustering based clinical characterization of the clusters identified by CIFU.

#### Circular Functional Modeling and Clustering

First, we introduce a method for clustering OCT HRC data into *K* homogeneous groups of samples (i.e., eyes). As an unsupervised approach, the method is based only on HRC data as input, and not any clinical variables of the samples. While the actual OCT measurements are taken on a discrete grid of angles around the center of ONH, in principle, these finely indexed measurements are assumed to vary continuously around a circular scale ranging from 0 to 360 degrees, and wrapped around. Thus, for statistical modeling, we will refer to the OCT data as a collection of *n* circular curves *X*_1_(*t*), *X*_2_(*t*) …, *X*_*n*_(*t*), where *X*_*i*_(*t*) represents the OCT value (of NRR thickness) in the *i*^*th*^ sample (*i = 1*, 2, … *n*) measured radially at each point *t* (here, *t* = 1,2, … 180) and around a common center. The angular indices are aligned for all samples and spaced 2 degrees apart starting from a common 0 degrees.

To allow for comparison of the curves based on their shapes rather than the magnitude, we normalize each curve *X*_i_ (*t*) by dividing it by 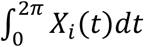.

Our modeling begins with the use of *p* basis functions for capturing the functional nature of data. If *ψ*_1_, *ψ*_2_, …, *ψ*_*p*_ are the basis functions with the associated basis expansion coefficients *γ*_*ij*_, where *i* = 1, 2, … *n* and *j* = 1, 2, … *p*, then the functional approximation for the *i*^*th*^ curve at point *t, X*_*i*_(*t*), is given by

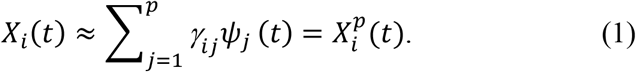

Our objective is to cluster each of the circular curves described as above into a pre-specified number (*K*) of clusters. Towards this, we used a discriminative functional mixture (DFM) model given by Bouveyron et al. (2015)^18^ in which *γ*_*i*_ *=* (*γ*_*i*1_, …, *γ*_*ip*_)^*t*^ of curve *X*_*i*_ follows a finite mixture model of *K* Gaussian components with the density function

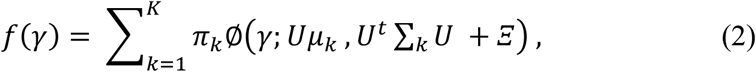

where π_*k*_ *≥* 0 is the mixing proportion of the *k*^*th*^ component (i.e., the cluster *k*) such that 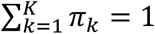, and ∅ is the standard Gaussian density function. Here, *U* is a *p* × *d* orthogonal matrix mapping the basis coefficients *γ* into the discriminative subspace (of dimension *d < p*) through a linear transformation. Similarly, *μ*_*k*_ and *Σ*_*k*_ are the mean vector and covariance matrix (for cluster *k*) of *γ* mapped into the discriminative subspace, and the noise of the above transformation is normally distributed with mean zero and covariance *Ξ*.

The DFM model was fit with an EM algorithm implemented in the R package funFEM^19^. Following the idea of constraining variance parameters in Fraley and Raftery (1999)^20^, the funFEM package allows 12 different choices of DFM models. As an initial step, using different restrictions on the noise covariance matrix *Ξ*, a preliminary model search was run with NRR data, with outlier curves included, over all 12 models and a flexible set of (61) basis functions. Based on the results of the initial run, and after removing the outlier curves by equation (3), the clustering algorithm was run using a smaller selective set of (21) basis functions.

A given curve *X*_*i*_(*t*) is considered an outlier if it exceeds the following threshold

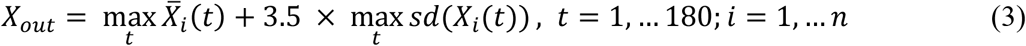

To avoid model overfitting, we determined the smallest number of basis functions (*p*) that recover the input curves sufficiently well, as determined by the fraction of variation explained (*FVE*) as described below. Let the sample mean of *n* given curves be

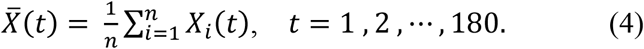

Then the total variation (TV) is given by

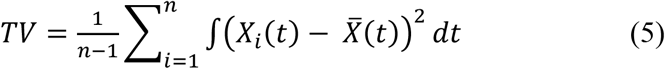

and the fraction of variation explained (FVE) by

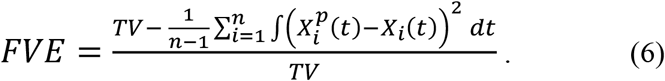

We used *FVE* as the criterion for selecting an optimal number of basis functions (*p*).

Finally, the number of clusters identified by the DFM models for each age group was determined by 3 known model selection criteria AIC, BIC and ICL.

For intuitive visualization of the clustering results, we plotted the curves of every cluster in a distinct color using a circular scale. The mean curve of each cluster, as computed by (4), is included as a bold black curve, which serves as a cluster-specific template.

For comparison of our circular functional clustering method with a popular non-circular data clustering approach, we used the Partitioning Around Medoids (PAM) as implemented in R. The optimal number of clusters identified by PAM was selected using the Average Silhouette Width (ASW), while Dunn Index was calculated as a measure of inter-cluster variation. We used the R packages ‘factoextra’ and ‘fpc’ for PAM clustering, validation and visualization.

#### Metaclustering and Clinical Characterization of Clusters

In the metaclustering step, the clusters identified by circular functional data were grouped based on their samples’ similarity in terms of a selected set of clinical variables that are known covariates of glaucoma. A feature selection step was performed simultaneously to detect the covariates that were the most distinctive across the metaclusters. The metaclustering workflow consists of the following steps:

1. In each age group, each circular functional cluster *C* was represented by the mean values of a set of clinical covariates of the samples in *C*.
2. We performed agglomerative hierarchical clustering of the clusters given by their mean covariate data with complete linkage, and a tuning parameter (wbound) to select the covariates that are the most distinctive across the metaclusters.
3. We plotted the metaclusters (identified by Step 2) with age group-specific dendrograms. A flat cut of the dendrograms at a common height threshold was use to distinguish the metaclusters in each age group, which were shown as subtrees of corresponding colors.
4. We visualized using contour plots the corresponding metaclusters of each age group to compare the distributions of the selected covariates across the metaclusters as well as the age groups.

The set of 9 covariates used in step 1, selected for their clinical relevance, are shown in Table 2. The R package ‘sparcl’^21^ was used for agglomerative and sparse hierarchical metaclustering in step 2; the arguments of the wbound parameter were varied from 2 to 5.

**Table 2:** The clinical covariates used for metaclustering in the three age groups. The two most significant covariates due to feature selection are shown in bold. The values of each variable in a metacluster are described as mean±sd. The units are given in parentheses. (*N*: the number of samples; IOP: IntraOcular Pressure; CCT: Central Corneal Thickness; CDR: Cup-to-Disc Ratio.)

|  | Age Group 1 |  |  | Age Group 2 |  | Age Group 3 |  |
| --- | --- | --- | --- | --- | --- | --- | --- |
| Metacluster {Clusters} | *M*<sub>1</sub> {3,6} | *M*<sub>2</sub> {1,2,5,7} | *M*<sub>3</sub> {4} | *M*<sub>1</sub> {1,5,7} | *M*<sub>2</sub> {2,3,4,6,8} | *M*<sub>1</sub> {3} | *M*<sub>2</sub> {1,2,4,5,6} |
| Metacluster *N* (percent) | 549 (29.8) | 1170 (63.6) | 122 (6.6) | 547 (40.5) | 804 (59.5) | 212 (27.1) | 569 (72.9) |
| IOP (mmHg) | 12.68±2.31 | 12.54±2.33 | 12.54±2.61 | 12.34±2.31 | 12.6±2.48 | 12.12±2.48 | 12.13±2.34 |
| CCT (μm) | 525.93±31.08 | 526.24±32.86 | 519.53±30.68 | 523.06±31.53 | 525.09±32.23 | 518.71±33.58 | 516.56±30.87 |
| Axial Length (mm) | 22.55±0.75 | 22.59±0.74 | 22.7±0.67 | 22.63±0.69 | 22.63±0.72 | 22.8±0.72 | 22.53±0.82 |
| RIM AREA (mm<sup>2</sup>) | 1.44±0.23 | 1.32±0.21 | 1.25±0.19 | 1.42±0.24 | 1.29±0.21 | 1.42±0.25 | 1.3±0.24 |
| DISC AREA (mm<sup>2</sup>) | 1.86±0.29 | 1.99±0.35 | 2.17±0.4 | 1.89±0.32 | 2.02±0.36 | 1.88±0.32 | 2.06±0.38 |
| DISC DIAMETER (mm) | 1.47±0.13 | 1.5±0.15 | 1.59±0.17 | 1.48±0.14 | 1.52±0.15 | 1.48±0.14 | 1.55±0.16 |
| VERTICAL CDR | 0.45±0.13 | 0.49±0.15 | 0.56±0.14 | 0.47±0.14 | 0.52±0.13 | 0.48±0.12 | 0.53±0.14 |
| **AVERAGE CDR** | **0.44±0.13** | **0.54±0.15** | **0.62±0.12** | **0.46±0.14** | **0.57±0.13** | **0.46±0.12** | **0.58±0.14** |
| **CUP VOLUME (mm<sup>3</sup>)** | **0.1±0.1** | **0.2±0.16** | **0.3±0.19** | **0.12±0.12** | **0.22±0.16** | **0.11±0.11** | **0.23±0.19** |

## 3. Results

The CIFU pipeline was run with HRC NRR phenotype data collected with OCT and clinical assessment of a normal cohort consisting of 3973 healthy eyes. The steps of the pipeline began with stratification of the OCT and clinical data by age into 3 age groups with (1) 1841, (2) 1351, and (3) 781 samples respectively. The list of clinical variables are summarized in Table 1. An identical sequence of steps of analysis was followed by CIFU within each age group.

**Table 1:**
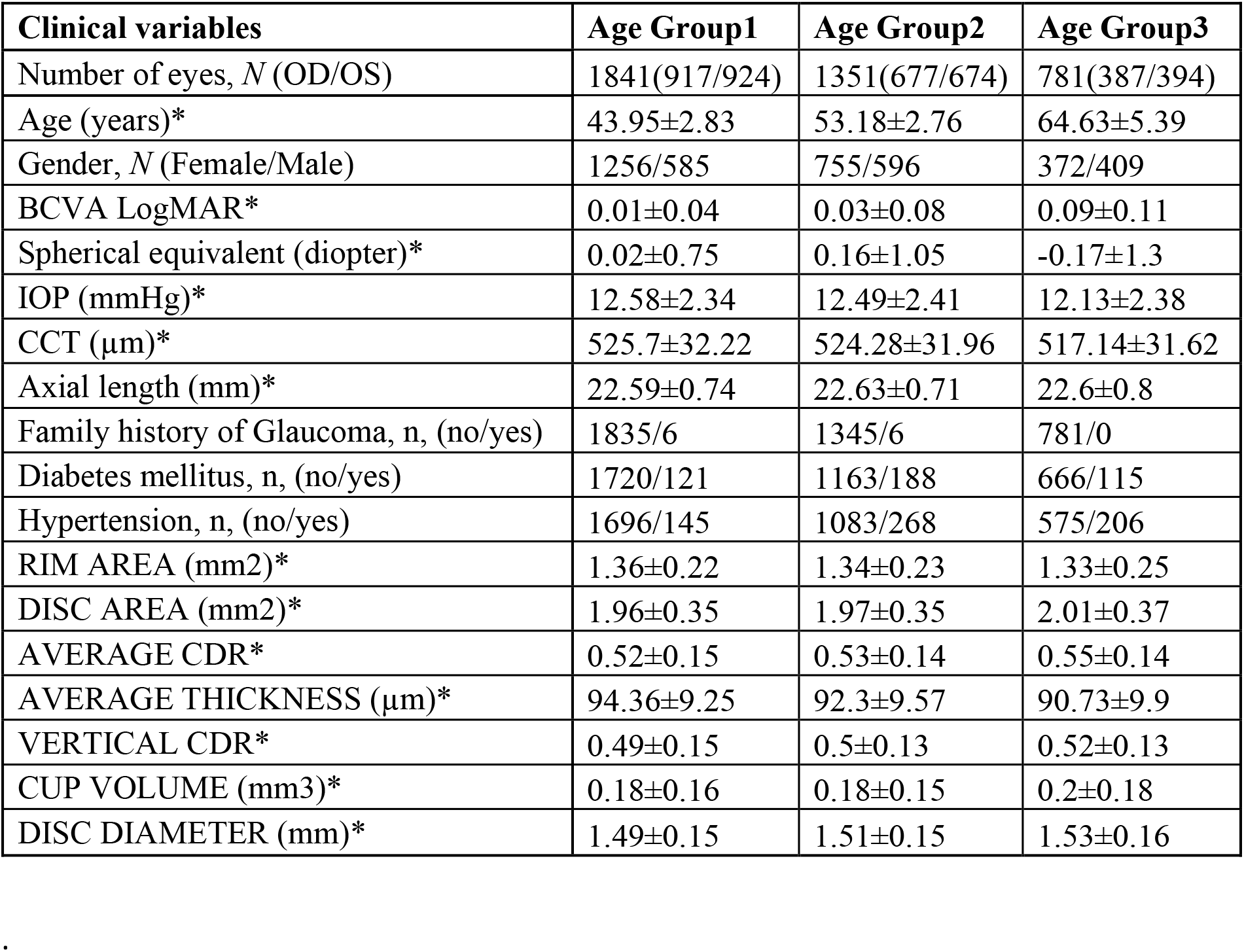
The clinical variables of the study participants in the three age groups. The units are given in parentheses. The asterisk(*) denotes that a variable is described as mean±sd. (*N*: Number of samples; OD: Oculus Dexter; OS: Oculus Sinister; BCVA LogMAR: Best Corrected Visual Acuity Logarithm of the Minimum Angle of Resolution; IOP: IntraOcular Pressure; CCT: Central Corneal Thickness; CDR: Cup-to-Disc Ratio.)

| Clinical variables | Age Group1 | Age Group2 | Age Group3 |
| --- | --- | --- | --- |
| Number of eyes, N (OD/OS) | 1841(917/924) | 1351(677/674) | 781(387/394) |
| Age (years)* | 43.95±2.83 | 53.18±2.76 | 64.63±5.39 |
| Gender, N (Female/Male) | 1256/585 | 755/596 | 372/409 |
| BCVA LogMAR* | 0.01±0.04 | 0.03±0.08 | 0.09±0.11 |
| Spherical equivalent (diopter)* | 0.02±0.75 | 0.16±1.05 | -0.17±1.3 |
| IOP (mmHg)* | 12.58±2.34 | 12.49±2.41 | 12.13±2.38 |
| CCT (μm)* | 525.7±32.22 | 524.28±31.96 | 517.14±31.62 |
| Axial length (mm)* | 22.59±0.74 | 22.63±0.71 | 22.6±0.8 |
| Family history of Glaucoma, n, (no/yes) | 1835/6 | 1345/6 | 781/0 |
| Diabetes mellitus, n, (no/yes) | 1720/121 | 1163/188 | 666/115 |
| Hypertension, n, (no/yes) | 1696/145 | 1083/268 | 575/206 |
| RIM AREA (mm<sup>2</sup>)* | 1.36±0.22 | 1.34±0.23 | 1.33±0.25 |
| DISC AREA (mm<sup>2</sup>)* | 1.96±0.35 | 1.97±0.35 | 2.01±0.37 |
| AVERAGE CDR* | 0.52±0.15 | 0.53±0.14 | 0.55±0.14 |
| AVERAGE THICKNESS (μm)* | 94.36±9.25 | 92.3±9.57 | 90.73±9.9 |
| VERTICAL CDR* | 0.49±0.15 | 0.5±0.13 | 0.52±0.13 |
| CUP VOLUME (mm<sup>3</sup>)* | 0.18±0.16 | 0.18±0.15 | 0.2±0.18 |
| DISC DIAMETER (mm)* | 1.49±0.15 | 1.51±0.15 | 1.53±0.16 |

The 180-point HRC data for each sample (eye) were modeled using *p* = 11 Fourier basis functions. We chose *p* = 11 since it was the smallest value of *p* for which *FVE*, as given in equation (6), exceeded 99% (Supplementary Figure S1). The curves were normalized and aligned to a common starting angle of 0 degrees to allow for comparison of their shapes around the center of ONH. Using equation (3), the outlier OCT samples were removed: 6 samples from age group 1, 1 from age group 2, and 5 from age group 3. Then, within each age group, the curves were clustered by a discriminative functional mixture (DFM) model as described in equation (2). The number of clusters (*K*) for each age group was determined by 3 different well-known criteria: AIC, BIC, and ICL. These criteria showed overall strong agreement attesting to optimal model selection as seen in Figure 1. Thus, we determined *K*, the number of clusters, for age group 1, 2, and 3 as 7, 8, and 6 respectively. The size of each cluster is shown in Table 2.

**Figure 1.**
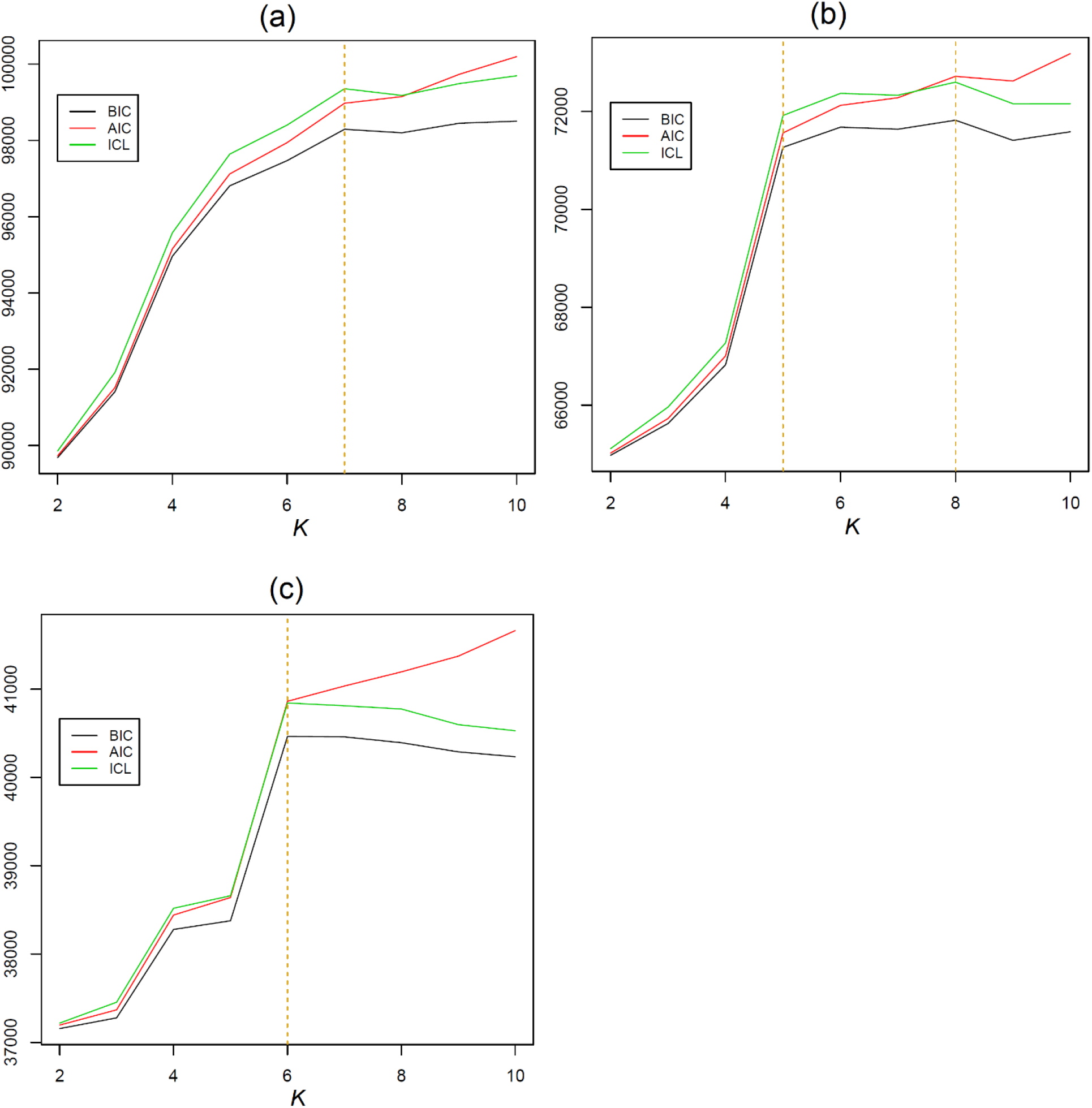
: The values of model selection criteria AIC, BIC, and ICL corresponding to fitting of a DFM model of *K* clusters to OCT NRR samples of age group 1 shown in (a), 2 in (b) and 3 in (c). The optimal DFM models for the age groups 1, 2 and 3 were selected for *K*=7, 8 and 6 respectively, beyond which no significant gain was noted.

The results of our circular functional clustering are shown in Figure 2(a)-(c) as a panel of *K* clusters for each age group. Each cluster *C* within a panel consists of the circular curves for the samples that belong to *C* (all shown in a common color specific to *C*) based on the similarity of their functional representation. To gain an intuitive understanding of the 180-point HRC OCT data on NRR phenotypes, we used a visualization of curves as represented on a common circular scale. Unsupervised clustering of the circular functions revealed various NRR patterns in the identified clusters, some of which were distinctive whereas others have subtle differences. Notably, the visualization reveals the unique mean shape (or NRR “template”) of each cluster as shown by a bold black circular curve in each plot of Figure 3.

**Figure 2.**
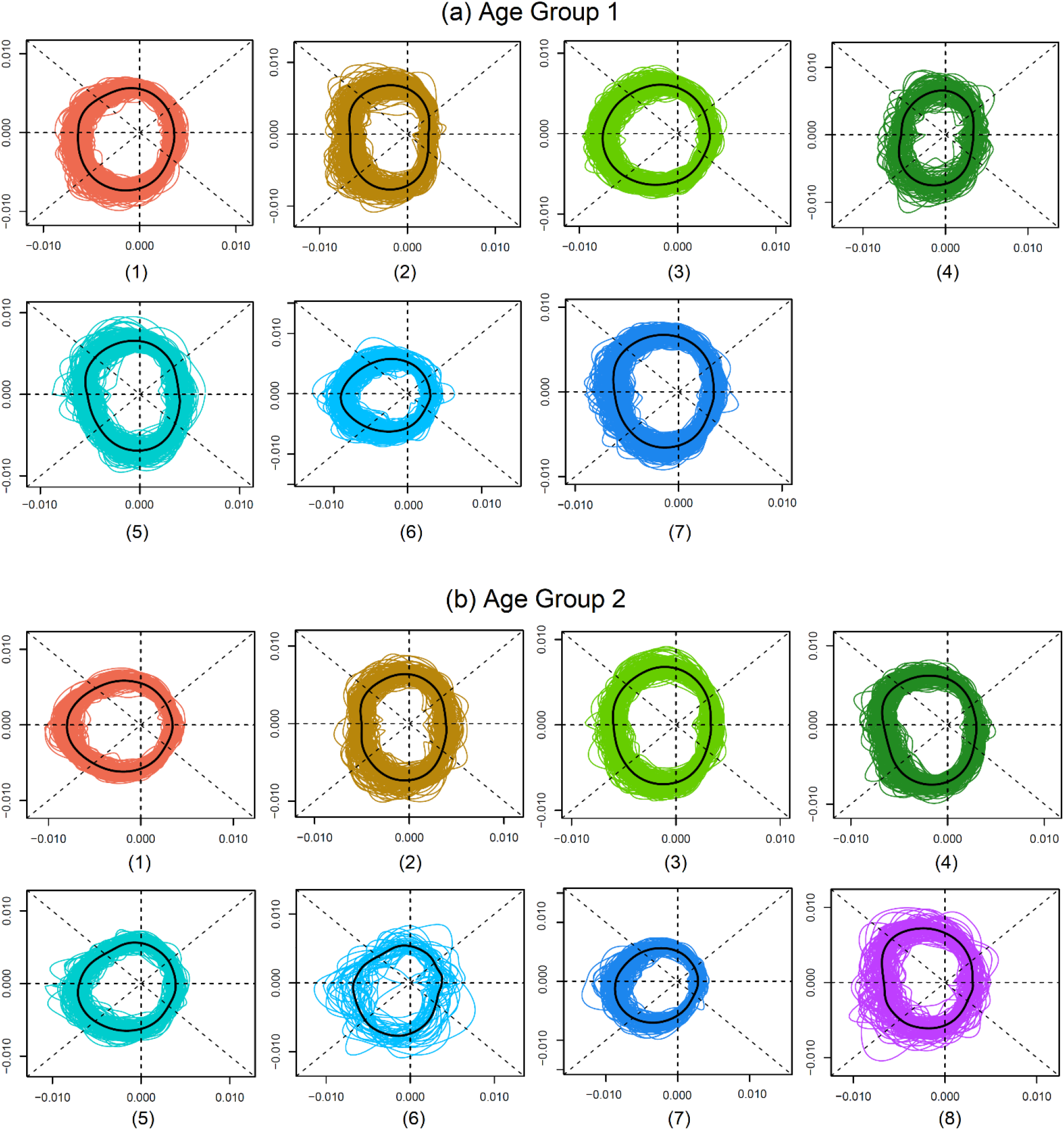
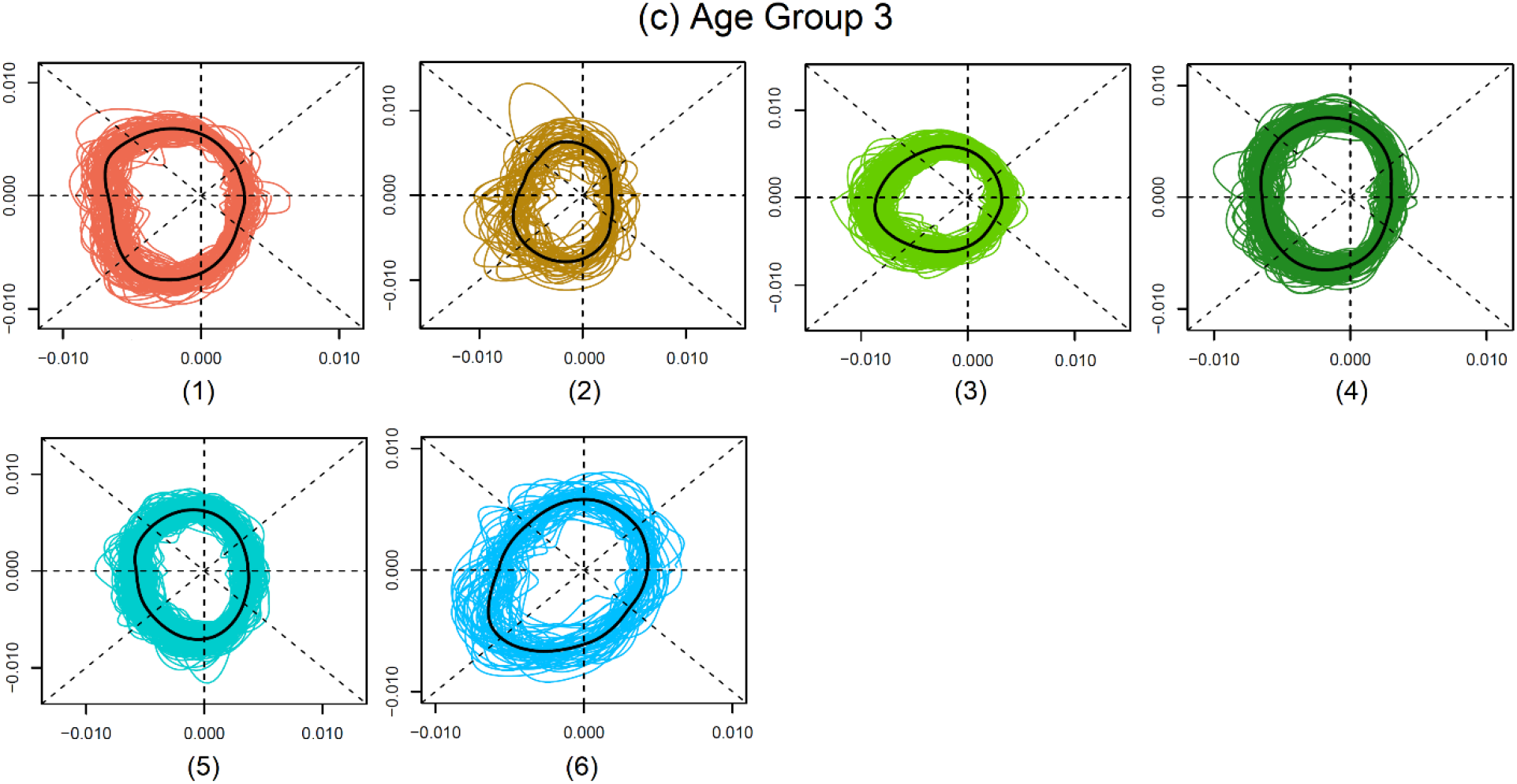
: The results of clustering the OCT NRR data of age groups 1, 2, and 3 are shown respectively in panels (a), (b), and (c) using a circular functional representation. The curves belonging to each cluster is shown in a distinct color within a panel. The sample mean of each cluster is shown as a bold black circular curve.

**Figure 3.**
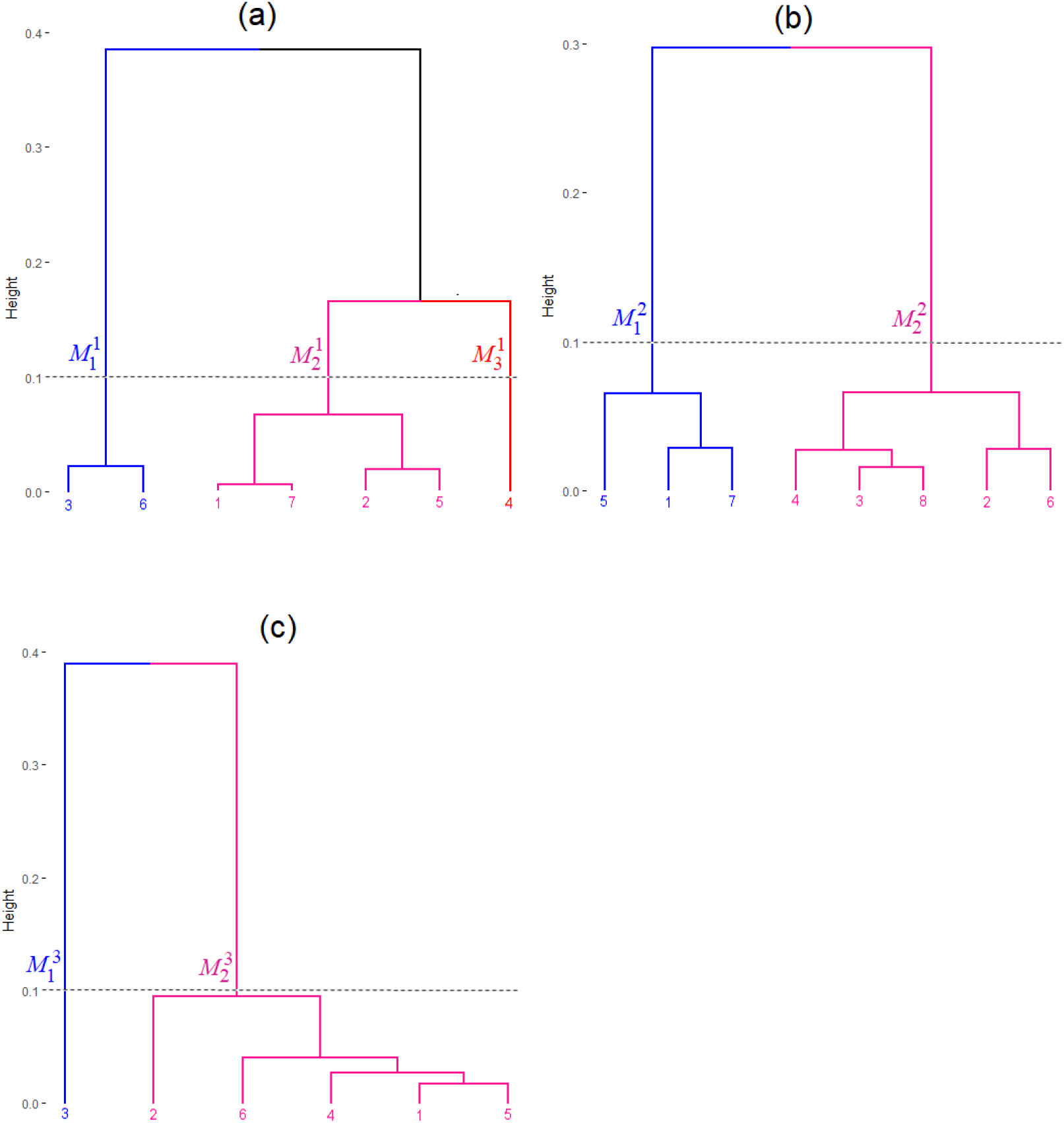
: The results of metaclustering are shown using dendrograms for age groups (a) 1, (b) 2, and (c) 3. The leaves denote the Id-s of the circular functional clusters of OCT NRR data. The metaclusters are obtained by a flat cut of each dendrogram at the common height of 0.1, and the labels and subtrees representing them are shown in different colors.

**Figure 4.**
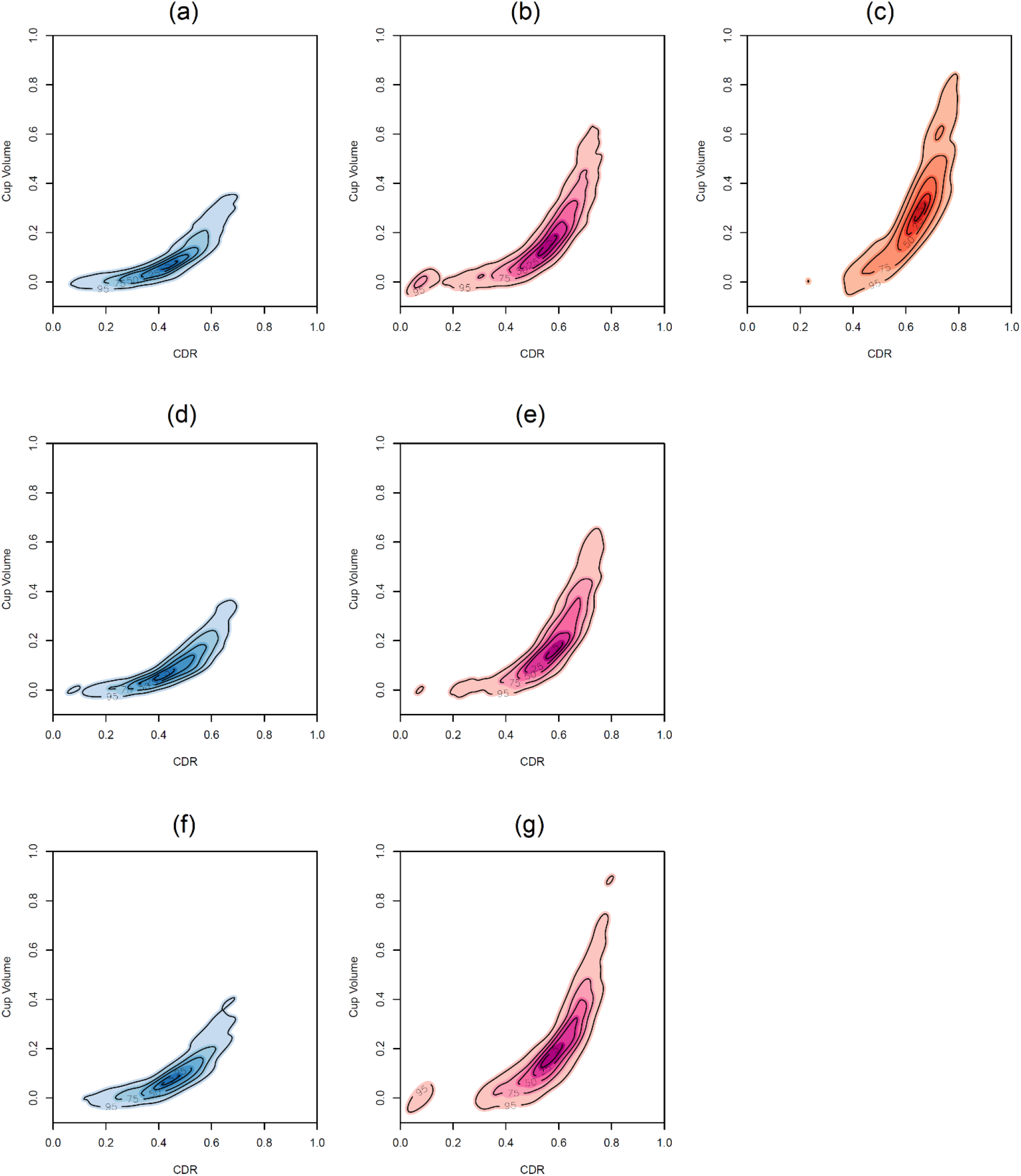
: Contour plots of the distributions of clinical covariates volume and average cup CDR of the samples belonging to metaclusters of age groups 1: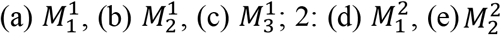; and 3: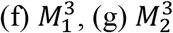. The corresponding metaclusters are shown in matched colors.

The circular curve visualization allows several interesting observations. We note the consistent dip at the temporal (T) region near 0 degrees, which is a characteristic feature shared by the templates of all clusters. This is supported by the well-known ISNT rule^22^ according to which, from the center of ONH, the rim is the thinnest at the temporal (T) region. Interestingly, we also observed various shapes and features in the cluster templates (such as distinctive protrusions, notches, tilts, etc.) that appear as well as vary continuously in different (non-T) regions around the circle. The clustering solution allows us to record the intra-cluster variation which could be used to quantitatively compare the dynamics (say, the rates of focal change) of corresponding clusters across age groups. In this regard, we note that non-circular (PAM) clustering of the same NRR HRC data yielded only 2 clusters of low phenotypic variation in every age group (Supplementary Figures S2 and S3) as noted by their small values of Dunn Index for every age group (1: 0.0548; 2: 0.044; 3: 0.0557).

In order to establish a correspondence among the clusters in different age groups as well as to characterize the samples that belong to each cluster, we conducted a metaclustering analysis. In this step, we clustered the clusters based on a set of 9 clinical variables of the samples therein. These are known covariates of glaucoma, and no NRR data from the previous clustering step was used. The results of sparse hierarchical metaclustering are shown in Figure 5. The dendrograms reveal the similarities among the clusters in terms of their mean sample covariates as well as the counts of metaclusters identified at different levels of each dendrogram. Based on flat cuts of all the dendrograms (at a common height threshold of 0.1), we identified 3 metaclusters 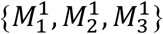, for age group 1; 2 metaclusters 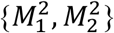 for age group 2; and 2 metaclusters 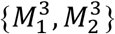 for agegroup 3. Notably, all the dendrograms show the metacluster 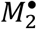 (pink subtree) to be more heterogeneous in every age group than the metacluster 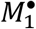 (blue subtree). Among the youngest participants, i.e., in age group 1, the metacluster 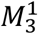 (consisting of the original cluster 4) is distinct from the metacluster 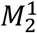.

A feature selection step, performed along with metaclustering, identified the optic disc cup volume and the average cup-to-disk ratio (CDR) of an eye as the most significant features in terms of the contributions of the different covariates to the metaclustering. These distinctive covariates allow us to register the correspondence of the metaclusters across the different age groups in Figure 6, which shows the contour plots of the metaclusters in their matched colors. The 3 metaclusters 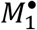 shown in blue have the smallest mean values of cup volume and CDR, the 3 metaclusters 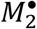 shown in pink have higher mean values of these covariates, and the single unmatched metacluster 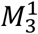 shown in red has the highest mean values (Table 2).

It is interesting to consider the unmatched metacluster 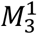 which not only has the highest mean values of the covariates (cup volume and average CDR) but is, in fact, comprised of a single, distinct cluster based on the OCT NRR phenotype data (Figure 5a). Here we note that notwithstanding a large value of CDR (especially *>* 0.*5*), cupping by itself is not indicative of glaucoma. In fact, it is known that deep but stable cupping can occur due to hereditary reasons without glaucoma (see Discussion). Rather, it is a change in these ONH parameters with age of the participants that is a clinical indicator of glaucoma. Since the samples included in the present study contain only healthy eyes as determined clinically by agreement of multiple glaucoma specialists, the presence of this unmatched cluster only in the youngest age group serves as a signature of healthy NRR phenotype with predictive potential for glaucoma. That is, the corresponding metacluster with such high values of these covariates among the older age groups would have the likelihood of progressing to glaucoma, and thus, is unlikely to be represented in a cohort of only healthy eyes, as we have in the present study.

## 4. Discussion

Unsupervised learning of the heterogeneity of normative ONH phenotypes in a given population can provide a more comprehensive understanding of the diversity of baselines that exist for degenerative neuropathies. Such knowledge is particularly useful in the case of glaucomas for which different ONH parameters play a combined role in early detection. For example, in a non-glaucoma multiethnic cohort of Asian individuals, the inter-eye RNFL profile was found by OCT to be less symmetric in Malays and Indians than that in Chinese^23^. Not only are the structural characteristics of individual eyes known to vary racially, even their rates of change over time could be different across population groups. For instance, the rate of change of BMO-MRW was recorded as *−*1.82 μm/year and *−*2.20 μm/year in glaucoma suspect eyes of European and African descents respectively^24^. In another multi-centred normal population study, both age-related decline and between-subject variability in BMO-MRW were observed^25^. Indeed, even the manufacturers of OCT technology noted racial differences in optic disc area, CDR, cup volume, and RNFL thickness when measured using their platform^26^.

The presence of phenotypic heterogeneity makes it less justified to apply common, universal thresholds for clinical determination of glaucomatous damage in different population groups using OCT measurements, particularly in the early stages of the disease when the baselines could have stronger initial effects. To account for the effects of normal variation in ONH parameters, large and racially representative normative databases of healthy eye OCT phenotypes should be created. However, often such collections of healthy samples tend to be small or moderately sized, e.g., the normative database of Cirrus HD-OCT platform included just 284 subjects^26^. In that cohort, Caucasians represented 43%, Chinese 24%, African Americans 18%, Hispanics 12%, and others 6%. The representation of Asian Indians, in contrast, was about 1% of the Cirrus HD-OCT cohort, which does not adequately reflect the 2020 projections about India to become the second in global glaucoma numbers, surpassing Europe^27^. Thus, more than 16 million Indians could be affected by glaucomas, and nearly 1.2 million get blind from the disease. Some resources such as the HRT3 Normative Database, while including 104 Indian individuals, did not improve the diagnostic sensitivity or specificity for glaucoma in that group for the potential reasons of limited sample size and intra-racial variation of ocular topography^28^.

In this study, we leverage on the large population-based LVPEI-GLEAMS study to generate new and relatively large OCT dataset based on nearly 4000 samples from normal Asian Indian participants. In fact, given that the recruitment of all the study participants was from a single geographic region (namely, the state of Andhra Pradesh), the scope of intra-racial variation to affect this dataset is limited. Moreover, the relatively large sample size of the data allowed us to identify a variety of clusters of NRR phenotypes, including the signature (cluster 4) with predictive potential for glaucoma consisting of 6.6% of all samples in the youngest age group (40-49 years). The absence of its corresponding cluster in the older healthy age groups despite their considerable sample sizes (total of 2132 samples of age 50 years and above) leads to a reasonable supposition of its potential pathological progression with increase in age, thereby resulting in lack of subsequent representation in a healthy cohort such as in the present study. Such phenotypic decline is consistent with the findings from a prospective longitudinal study that found the rate of age-related, glaucomatous global (or global percentage) rim area loss to be 3.7 (5.4) times faster as compared to healthy eyes^29^.

Importantly, independent support for the identified signature relies on its clinical characterization in terms of covariates such as cup volume and CDR, which are useful parameters for diagnosis of glaucoma suspect^30^. Despite its normal mean value of intraocular pressure (IOP) as is expected of healthy eyes, the signature cluster has the highest mean values of average CDR and cup volume of all metaclusters across all 3 age groups (Table 2). In a recently published longitudinal study that started from baseline values and was run over a 5-year period, the covariates which had statistically significant increase in glaucomatous progression included CDR and cup volume^31^. We understand that it would perhaps be ideal to follow-up healthy individuals and measure the changes in their clinical covariates as they age in order to classify the ONH phenotypes via supervised learning. The approach of CIFU, in comparison, involves unsupervised learning of different high-resolution phenotypes in age-stratified data and their characterization using key covariates, which is far less time-consuming and yet has the potential to produce a clinically insightful database for diverse populations with high phenotypic heterogeneity.

In addition to its sample size and racial representation, perhaps the most noteworthy feature of the present OCT dataset is its unique HRC measurements of NRR thickness. These samples, collected at every 2 degrees, extend the typical use of such measurements recorded at either 4 quadrants or 12 clock-hours, and even 48 angular positions^32^, to higher-dimensional analysis. As clustering with curves show, the focal variations could be more nuanced than that suggested by a general rule, e.g., ISNT, and a capability to “zoom” into finer angular divisions can reveal further patterns^33^. In low-resolution data, it may be difficult to detect focal changes within the confines of pre-determined inflexible sectors. Moreover, the templates of the clusters could also be compared using known tests in shape analysis^34^. Be it circular data from OCT, or optic phenotypes in general, they seem suitable as candidate applications of circular statistics, and yet, we are unaware of any major previous studies in this regard. Further, the high-resolution also allowed the HRC data to be closely approximated by continuous curves, and thus, specified by the corresponding functional representation. While clustering of circular point data^12,35,36^ as well as clustering of curves^37–39^ and (non-circular) functional data^13,14,19,40–42^ have been addressed by past studies, the present clustering of curves in the form of circular functions is possibly a novel application.

We understand that the present study has certain limitations. As we noted above, a prospective cohort study would be better suited to validate the predictive glaucomatous potential of the identified NRR phenotypic signature. We plan to address this in our future work. While HRC data could be accessed from the OCT scans, it is not commonly done by the clinical protocols. We hope that by adding user-friendly interfaces to CIFU, we may promote such data acquisition and analysis. Indeed, there are several distinct advantages of our approach which could be built upon further in future studies. The numeric representation of the functional mixture model parameters could be used to compare the normal against disease ONH phenotypes allowing us to characterize any changes therein with precision and rigor. Once a database of phenotypic parameters is developed, known measures of shapes and distances between curves could be used for objective clinical classification of new samples. Applied to longitudinal analyses, our high-resolution modeling could identify intermediate, or previously uncharacterized, stages of disease progression, especially by focusing on variations within fine angular sections. Straightforward extensions are feasible for similar circular data such as RNFL phenotypes and other optic neuropathies as well as related eye imaging platforms, e.g., OCT-Angiography (OCTA). As we have demonstrated for other biomedical platforms^39,43–46^, the new pipeline CIFU could be enhanced incrementally with different functionalities, say, to increase computational efficiency or capture the perspective of the clinical experts. The circular curve visualization introduced in the present study may lead to a more user-friendly tool for clinical purposes as we plan to make it interactive, with advanced capabilities to jointly handle data and metadata, in our future work.

## Data Availability

The de-identified data are available upon request from the authors.

## Acknowledgements

AP is supported by NSF grant DMS-1811888. The population-based cohort was funded by Hyderabad Eye Research institute.

## Supplementary Figures

**Supplementary Figure S1:**
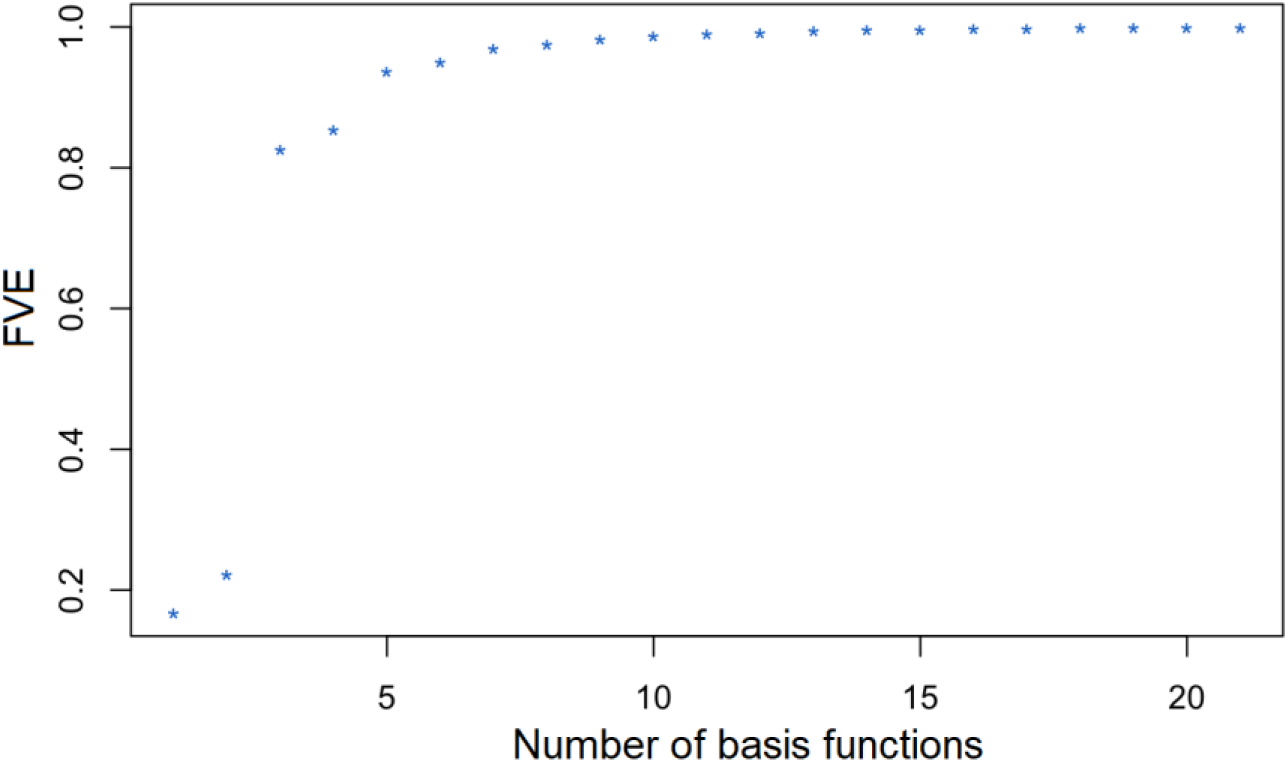
The fraction of variation explained (*FVE*) by models with different choices of the number (*p*) of basis functions used for the functional representation of OCT data.

**Supplementary Figure S2:**
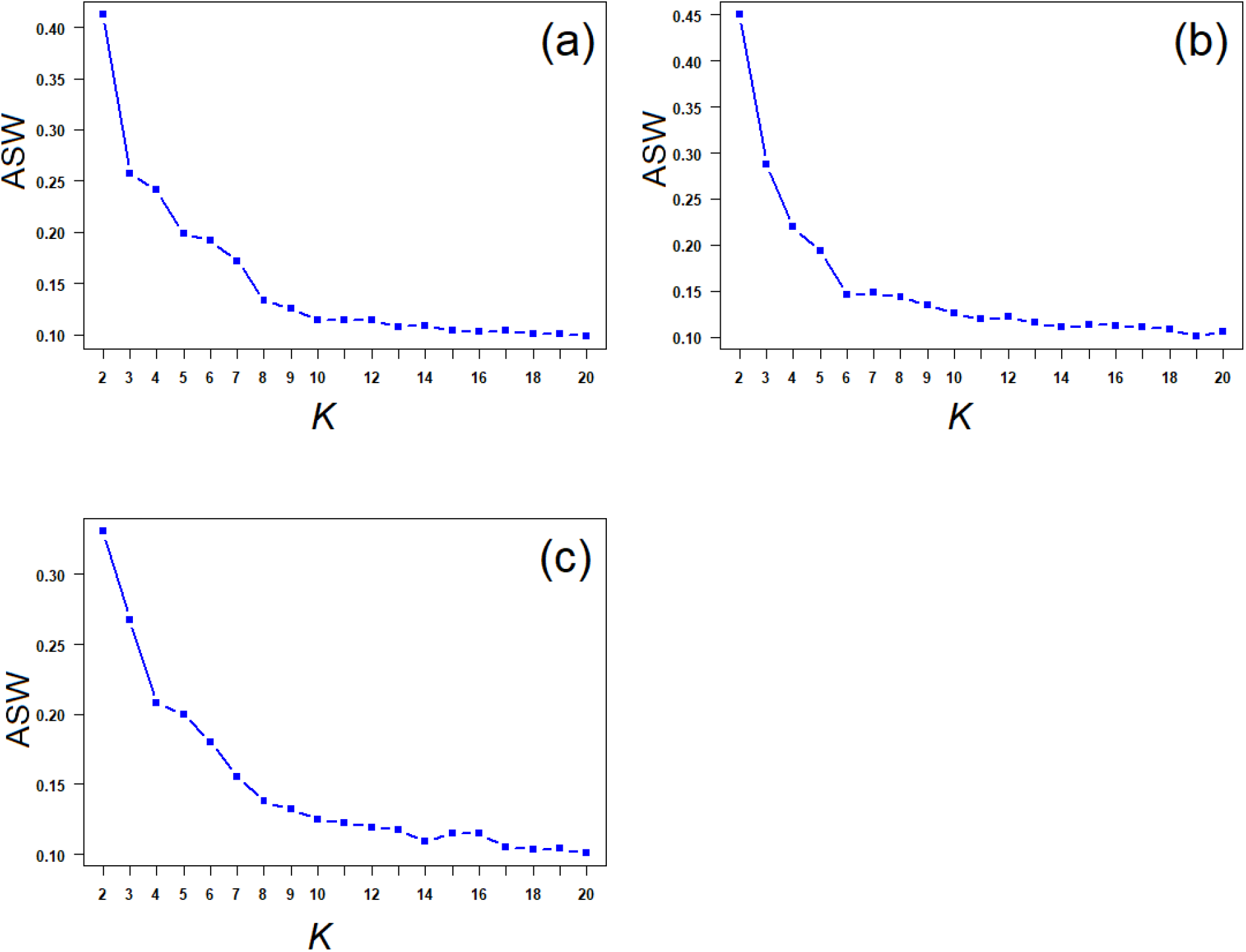
Average Silhouette Width (ASW) for different choices of the number of clusters (*K*) due to PAM clustering of OCT HRC data for age groups (a) 1, (b) 2, and (c) 3. The value of ASW is maximum for *K*=2 for all three age groups.

**Supplementary Figure S3:**
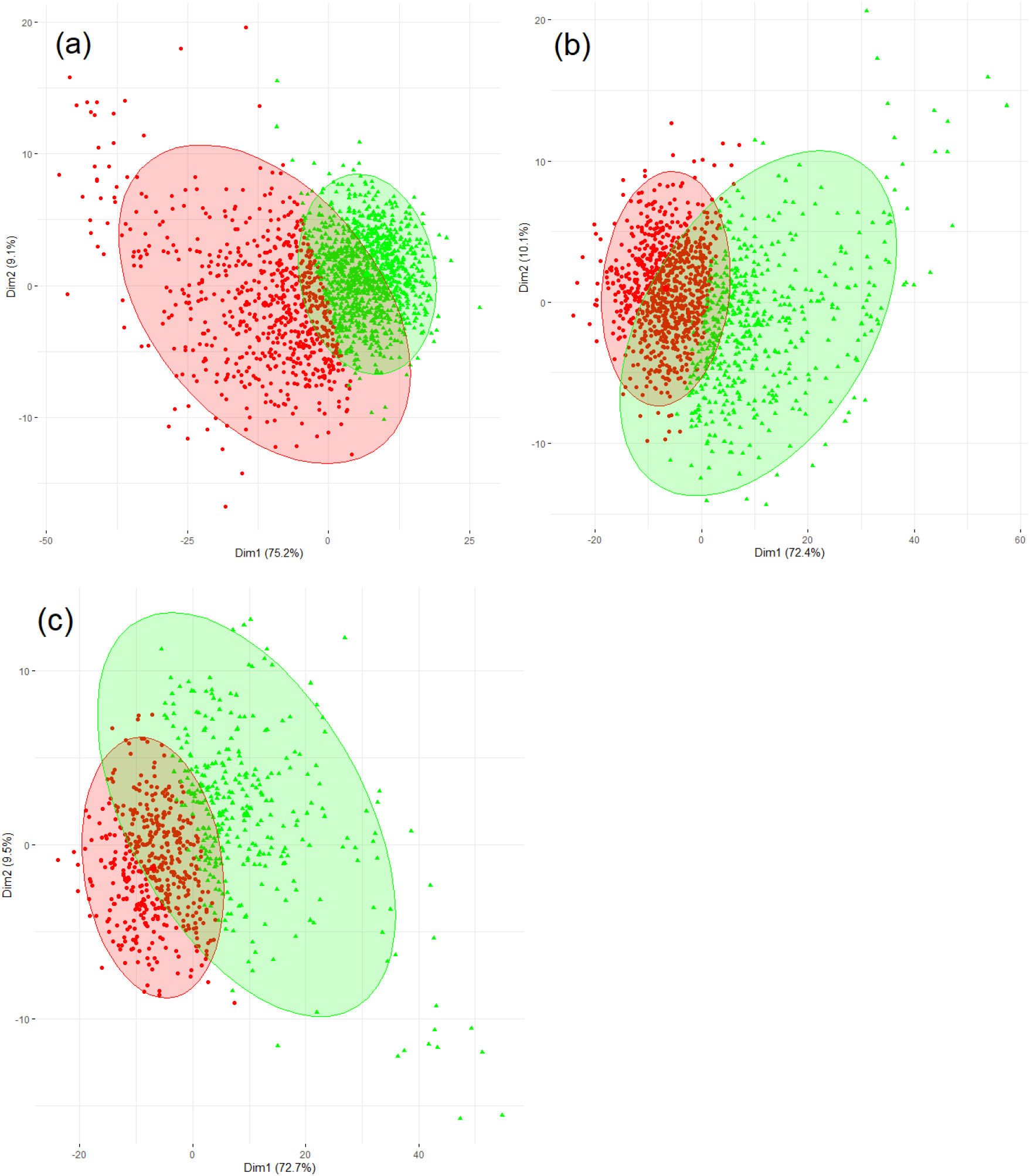
Results of PAM clustering OCT HRC data for age groups (a) 1, (b) 2, and (c) 3. Each scatterplot shows *K*=2 clusters (in red and green) as identified by PAM based on maximum ASW. Each 2D point represents a sample’s first two principal components of HRC data.

## Notes

### Competing Interest Statement

The authors have declared no competing interest.

### Funding Statement

No external funding was received to conduct this study.

### Author Declarations

"Written informed consent was obtained from all participants to participate in the study, and the ethics and review committee of the LVPEI reviewed and approved the methodology and was conducted in strict adherence to the tenets of the Declaration of Helsinki."

